# RSV-Related Healthcare Burden: A Prospective Observational Study in a Resource-Constrained Setting

**DOI:** 10.1101/2024.04.29.24306491

**Authors:** Senjuti Saha, SM Sudipta Saha, Naito Kanon, Yogesh Hooda, Mohammad Shahidul Islam, Shuborno Islam, Zabed Bin Ahmed, Sheikh Wasik Rahman, Md Jahangir Alam, Ataul Mustufa Anik, Probir K Sarkar, Mohammed Rizwanul Ahsan, Md. Ruhul Amin, Samir K Saha

**Affiliations:** Child Health Research Foundation, Dhaka, Bangladesh; Department of Social and Behavioural Sciences, Harvard T. H. Chan School of Public Health, Boston, USA; Department of Pediatric Respiratory Medicine (Pulmonology), Bangladesh Shishu Hospital and Institute, Dhaka, Bangladesh; Department of Emergency, Observatory and Referral, Bangladesh Shishu Hospital and Institute, Dhaka, Bangladesh; Department of Microbiology, Bangladesh Shishu Hospital and Institute, Dhaka, Bangladesh

**Author notes:** Correspondence to Senjuti Saha, PhD Deputy Executive Director Child Health Research Foundation 23/2 Khilji Road, Mohammadpur, Dhaka 1207, Bangladesh And Professor Samir K Saha, PhD, FAAM, FRCPath Executive Director Child Health Research Foundation, 23/2 Khilji Road, Mohammadpur Dhaka 1207, Bangladesh Head, Department of Microbiology, Bangladesh Shishu Hospital & Institute Sher-e-Bangla Nagar, Dhaka 1207, Bangladesh.

## Abstract

**Background:** Respiratory syncytial virus (RSV) is a leading cause of pediatric hospitalizations worldwide, straining health systems. Data gaps in resource-limited settings and limited RSV immunization impact estimates hinder policymaking.

**Methods:** From January to December 2019, we conducted a prospective study at Bangladesh’s largest pediatric hospital to assess RSV’s burden on the health system. Outcomes for RSV-positive under-five children were analyzed. We followed outcomes of children denied hospitalization due to bed shortages. Adjusted hazard ratios for children denied admission versus admitted were estimated using survival analysis. Monte Carlo simulations with a queuing model were used to estimate effects of RSV interventions on admission denials and mortality.

**Findings:** Of 40,664 children admitted, 31,692 were under-five; 19,940 were in study wards. Among 7,191 admitted with possible respiratory infections, 6,149 (86%) had samples taken, with 1,261 (21%) testing RSV-positive. Median age of RSV cases was 3 months, with a median hospital-stay of 5 days; 24 (1.9%) died in hospital. RSV cases accounted for 8,274 of 151,110 bed days. Additionally, of 9,169 children denied admission, outcomes were tracked for 3,928 and compared with 2,845 admitted. The hazard ratio for death was 1.56 [95%CI:1.34-1.81] for children denied versus admitted, highest within neonates at 2.27 [95%CI:1.87-2.75]. RSVpreF maternal vaccine or Nirsevimab could have reduced denials by 677 [95%PI:63-1347] and 1,289 [95%PI:684-1865], respectively, potentially preventing 130 [95%PI:-60-322] and 258 [95%PI:32-469] deaths.

**Interpretation:** RSV strains healthcare in Bangladesh, increasing mortality risks. Preventive interventions could lessen its impact, boosting healthcare capacity and child health in resource-limited settings.

## Introduction

Respiratory syncytial virus (RSV) is the leading cause of acute lower respiratory infection in young children. Annually, RSV infections are estimated to result in 33 million episodes, 3.6 million hospitalizations, and over 100,000 deaths worldwide.^1^ Low- and middle-income countries (LMICs) bear a disproportionate burden of the disease, where over 95% of RSV-associated acute lower respiratory infection episodes and more than 97% of RSV-attributable deaths occur.^1^

Interventions, such as maternal vaccines and long-acting monoclonal antibodies, promise to reduce RSV-related morbidity and mortality.^2–6^ There is also growing evidence suggesting that these interventions might have broader benefits, like reducing respiratory illnesses caused by other pathogens, and decreasing recurrent hospitalizations.^7–9^ Policymakers are grappling with how to prioritize and implement these measures, but often lack up-to-date and locally relevant epidemiological data from LMICs to guide these decisions.

In Bangladesh, a densely populated lower-middle income country, with a birth cohort of almost 3 million, the healthcare system faces challenges due to the high burden of communicable and non-communicable diseases.^10,11^ Data from the country’s largest pediatric hospital indicate that approximately 20% of children needing hospitalization—whether for communicable or non-communicable diseases—have to be turned away due to bed shortages.^12^ In these settings, a high RSV burden can exacerbate capacity issues while also potentially leading to adverse health outcomes for children who are denied admission. Thus, it is imperative to consider both the direct and indirect effects of RSV infections on child health and the broader health system when making informed policy decisions.

Two multi-country studies, Aetiology of Neonatal Infections in South Asia (ANISA),^13^ and Pneumonia Etiology Research for Child Health (PERCH),^14^ have highlighted RSV as a primary etiological agent in sepsis and respiratory tract infections in Bangladesh and other countries in Asia and Africa. While these studies underscore RSV’s role in child morbidity in resource-constrained settings, gaps remain in understanding the broader impact of RSV-associated hospitalizations on the healthcare capacity in resource-constrained settings.

To address the gaps in data and estimate the potential impact of RSV vaccines, we conducted a prospective, observational study in Bangladesh’s largest pediatric hospital. We aimed to quantify RSV-associated hospitalizations, bed usage, and outcomes. We also aimed to assess the broader health implications for children denied admission due to bed shortages by simulating different scenarios involving RSV preventive interventions, providing crucial data to inform policy decisions.

## Methods

### Ethical considerations

This study was approved by the ethical review board of the Bangladesh Shishu Hospital and Institute (BSHI). For children admitted to the hospital, written consent was obtained from parents or guardians for nasopharyngeal sample collection, testing and inclusion in the study. For children denied hospitalization, data on their health outcome of children were obtained from caregivers over telephone after obtaining verbal consent.

### Study Design and Setting

From January to December 2019, we conducted a prospective observational surveillance study at BSHI, located in Dhaka, Bangladesh. BSHI, with its 653-bed capacity (2019), is Bangladesh’s largest pediatric hospital, offering primary to tertiary care for patients up to 18 years old. Admission decisions for the in-patient department (IPD) are made by physicians in the Emergency Room (ER).

This study had two main components: (i) active surveillance within the IPD to detect RSV infections; and (ii) active surveillance of health outcomes of patients who, despite needing hospitalization, were denied admission due to bed shortages in the IPD, and a control group of patients who were admitted to the IPD.

### RSV Surveillance in the IPD

Children aged 0–59 months admitted to the wards selected for the study were evaluated by study physicians. Inclusion criteria for this study were children hospitalized with a possible respiratory infection as guided by the WHO RSV hospital-based surveillance case definition (Text S1, appendix page 3).^15^ These study wards contained 414 beds; the remaining 239 beds of the total 653 were not included because they were in wards that do not admit patients with possible infectious diseases (surgery, nephrology, oncology and cardiology wards). Nasopharyngeal swabs were collected by trained nurses from eligible children except those requiring support such as high-flow nasal cannula, CPAP, headbox, or incubators (Table S2, appendix page 24), and tested using qPCR (Text S2, appendix page 4).

### Surveillance of Children Denied Admission in the ER

Upon arrival at BSHI, families with children potentially needing hospitalization are triaged in the ER. Beds are available on a first-come first-serve basis. If the physician decides that the child requires hospitalization, but no beds are available, families are referred or advised to seek care elsewhere. During this study, when a child could not be admitted due to bed shortages despite requiring hospitalization (referred to here as “denied”), trained research assistants documented the ER physician’s diagnosis for the patients and collected the parents’ or a family member’s contact information during their hospital visit (between 8 am and 9 pm) and obtained permission to call them later (Text S1). Here, “denials” refer only to denial at BSHI and do not indicate that children were unable to receive care at a different hospital.

### Health Outcome Follow-Up

Among the under-five children denied admission due to bed shortages and whose contact information was obtained, families were selected using a computer-based randomization algorithm and contacted via telephone approximately two weeks after denial of admission, and the child’s health status (“alive” or “deceased”), and date of death (if applicable) were recorded upon obtaining verbal consent on study participation and data collection. If the child was alive during the 2-week follow-up, a second follow-up call was made three months after denial of admission. In parallel, from the cohort of under-five children who were admitted during the same time, children were randomly chosen for follow-up in the same manner.

Survival curves were computed using the Kaplan-Meier method and crude and adjusted hazard ratios were calculated using Cox Proportional-Hazards models (Text S3).

### Estimated impact of RSV preventive interventions

We used queueing theory^16^ and Monte Carlo simulations to understand how a reduction in RSV cases requiring admission may impact: (i) strain on hospital capacity, (ii) overall denial of patients seeking admission, and (iii) mortality. Our model algorithm was based on the admissions process described above and simulated daily admissions and discharges through the hospital given observed and hypothetical caseloads. Details of the simulations are provided in the appendix (Text S4, Figures S1, S2).

## Results

### Cases Hospitalized with RSV

In 2019, a total of 49,833 children sought care at BSHI and were advised admission, of whom 40,664 could be admitted based on the available beds (Figure 1, Table 1). 31,692 of them were <5 years old, and of them 19,940 were admitted in the screening wards. Of those admitted, 7,191 met the RSV hospital-based surveillance case definition, and a sample was successfully collected from 6,149 (86%) of these children. 1,042 children were not enrolled because 152 did not consent, 669 left before NP swabs could be collected, and in 221 cases sample collection was not possible for various reasons such as use of high-flow oxygen canula or being in a special ward (Figure 1, Table S2). 92% (5658/6149) of all samples were collected within 72 hours of hospitalization (Table S3).

**Figure 1.**
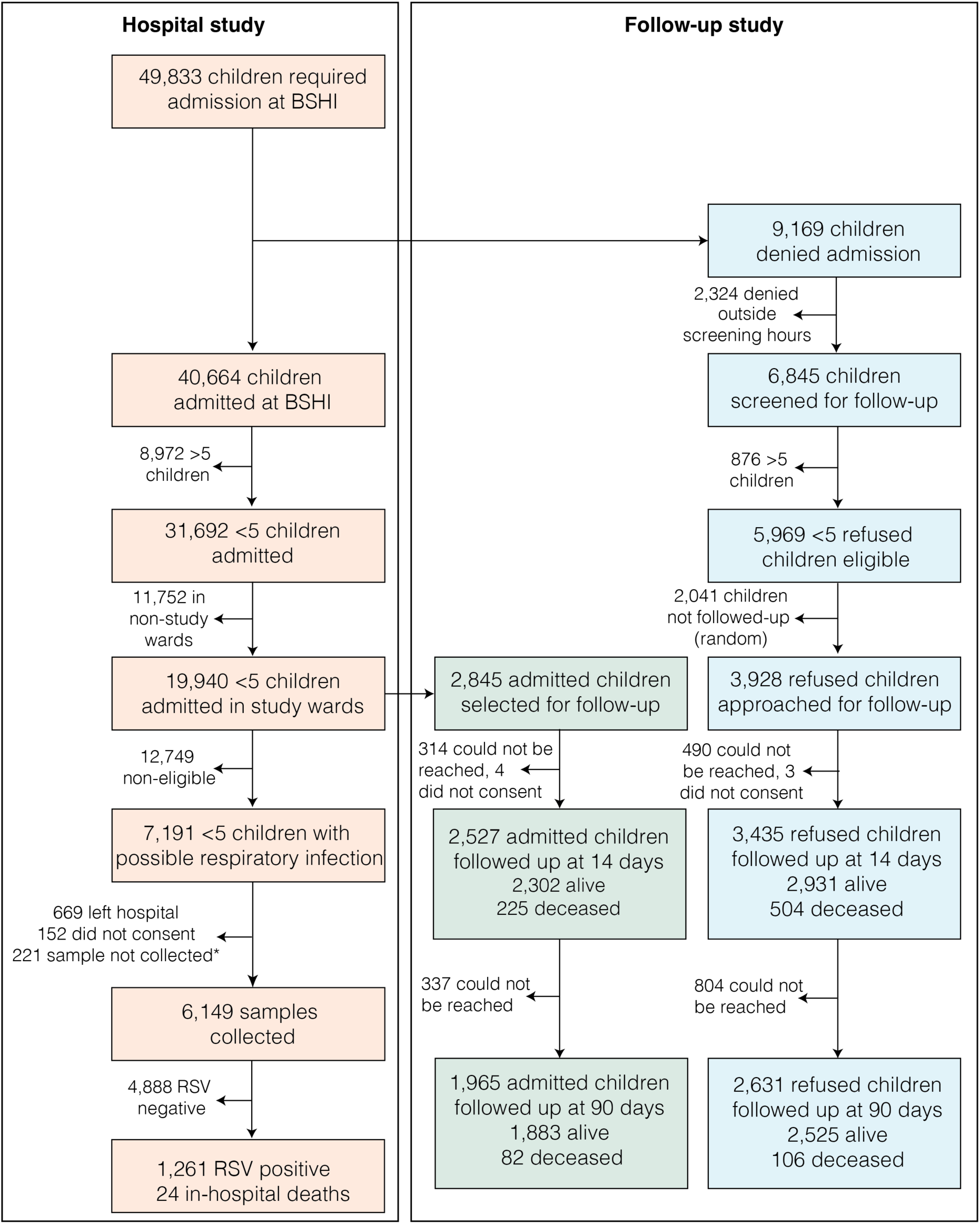
Flowchart of sample collection and patient follow-up. The figure illustrates the process of sample collection and subsequent patient follow-up for children under five admitted to BSHI or denied admission due to the unavailability of beds. *Please see appendix Table S2 for reasons why samples could not be collected.

**Table 1:** Clinical attributes of <5-year children admitted to study wards by RSV status. p-values were computed with Chi-squared tests for categorical variables (sex, case fatality rate, diagnosis), and Kruskal-Wallis tests for the continuous variables (age, length-of-stay). *A list of diagnoses that may be included in “other” is provided in the Appendix, Table S1.

|  | Overall | RSV Positive | RSV Negative | Eligible but untested | Not eligible | p-value |
| --- | --- | --- | --- | --- | --- | --- |
| **Number of Patients** | 19940 | 1261 | 4888 | 1042 | 12749 |  |
| **Sex - Female (%)** | 7582 (38.0) | 386 (30.6) | 1755 (35.9) | 392 (37.6) | 5049 (39.6) | <0.0001 |
| **Age in months (median [IQR])** | 5.00 [0.00, 16.00] | 3.00 [1.00, 8.00] | 2.00 [0.00, 10.00] | 0.00 [0.00, 6.00] | 7.00 [0.00, 21.00] | <0.0001 |
| **< 2 weeks** | 5448 (27.3) | 72 ( 5.7) | 1486 (30.4) | 466 (44.7) | 3424 (26.9) |  |
| **2 weeks - 3 months** | 3955 (19.8) | 625 (49.6) | 1214 (24.8) | 262 (25.1) | 1854 (14.5) |  |
| **4 - 6 months** | 1648 ( 8.3) | 184 (14.6) | 500 (10.2) | 78 ( 7.5) | 886 ( 6.9) |  |
| **7 months - 2 yrs** | 5693 (28.6) | 312 (24.7) | 1194 (24.4) | 185 (17.8) | 4002 (31.4) |  |
| **2 – 5 yrs** | 3196 (16.0) | 68 ( 5.4) | 494 (10.1) | 51 ( 4.9) | 2583 (20.3) |  |
| **In-hospital case fatality (%)** | 1266 (6.3) | 24 (1.9) | 372 (7.6) | 291 (27.9) | 579 (4.5) | <0.0001 |
| **Length of stay (median [IQR])** | 4.00 [2.00, 7.00] | 5.00 [4.00, 8.00] | 6.00 [3.00, 9.00] | 2.00 [1.00, 4.00] | 4.00 [2.00, 7.00] | <0.0001 |
| **Diagnosis Clusters (%)** |  |  |  |  |  |  |
| **Respiratory Manifestation** | 5476 (27.5) | 988 (78.4) | 2392(48.9) | 475 (45.6) | 1621 (12.7) | <0.0001 |
| **Gastrointestinal Manifestation** | 2498 (12.5) | 45 ( 3.6) | 170 (3.5) | 24 ( 2.3) | 2259 (17.7) | <0.0001 |
| **Perinatal Asphyxia** | 2570(12.9) | 24 ( 1.9) | 936 (19.1) | 285 (27.4) | 1325( 10.4) | <0.0001 |
| **Febrile Illness** | 1548( 7.8) | 25 ( 2.0) | 209 ( 4.3) | 24 ( 2.3) | 1290 ( 10.1) | <0.0001 |
| **Preterm low-birth weight** | 1136 ( 5.7) | 19 ( 1.5) | 341 ( 7.0) | 127 ( 12.2) | 649 ( 5.1) | <0.0001 |
| **Systemic Infections** | 2782( 14.0) | 131 ( 10.4) | 831 ( 17.0) | 269 ( 25.8) | 1551 ( 12.2) | <0.0001 |
| **Other\*** | 12817 (64.3) | 466 (37.0) | 3064 (62.7) | 643 (61.7) | 8644 (67.8) | <0.0001 |

All 6,149 samples were tested for the presence of RSV using qPCR, and of them 1,261 (21%) samples tested positive. Assuming 1,042 untested eligible cases among 7,191 had the same positivity as those tested, the overall proportion of all hospital admissions that were RSV positive was 4.65% or 465 RSV cases per 10,000 <5 hospitalizations (95%CI:443-489).

Over 70% (882/1261) of the total RSV-positive cases were enrolled in the study during the four months from June to September (Figure 2B). During July, August and September, more than 20% of the 414 beds were occupied by RSV-positive cases (Figure 2C).

**Figure 2.**
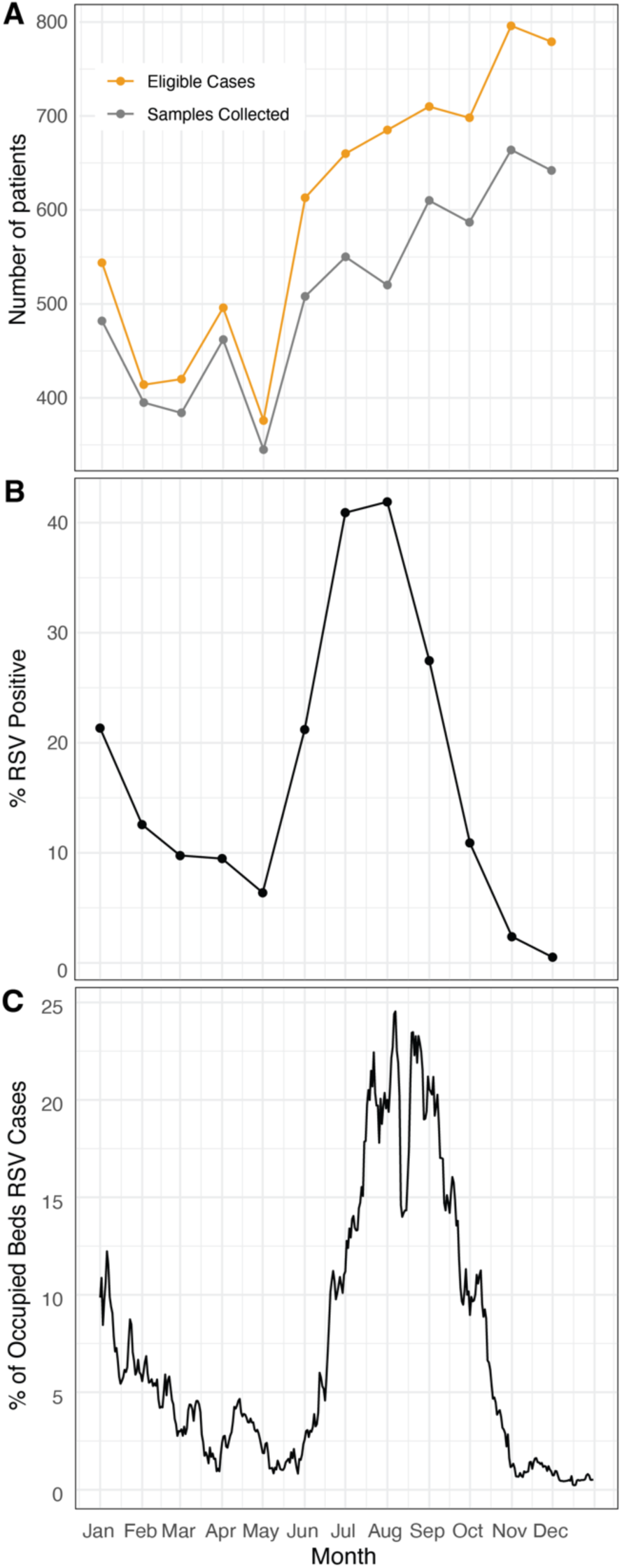
RSV Infections and Bed Occupancy in BSHI in 2019. A. The total number of patients meeting the eligibility criteria of the study each month, and the number of samples collected from these patients. B. Percentage of tested cases that were RSV-positive each month. C. Percentage of occupied beds with RSV-positive cases across different months. The dip in bed occupancy seen in mid-August is likely because of the Eid-ul-Adha holidays.

Of the positive cases, 30.6% were females. The median age of RSV-positive cases was 3 months (IQR:1.0-8.0) and 70% (881 of 1261) of the cases were within their first six months of life (Figure 3A, Table 1). 17% of infections occurred in the first four weeks of life, highlighting the vulnerability of very young babies to RSV. Median hospital stay of RSV cases was 5 days (IQR: 4-8), while median stay of all hospitalized cases was 4 days (IQR:2-7). Throughout the year, RSV cases accounted for 8,274 (5.5%) of the total 151,110 bed days observed.

**Figure 3.**
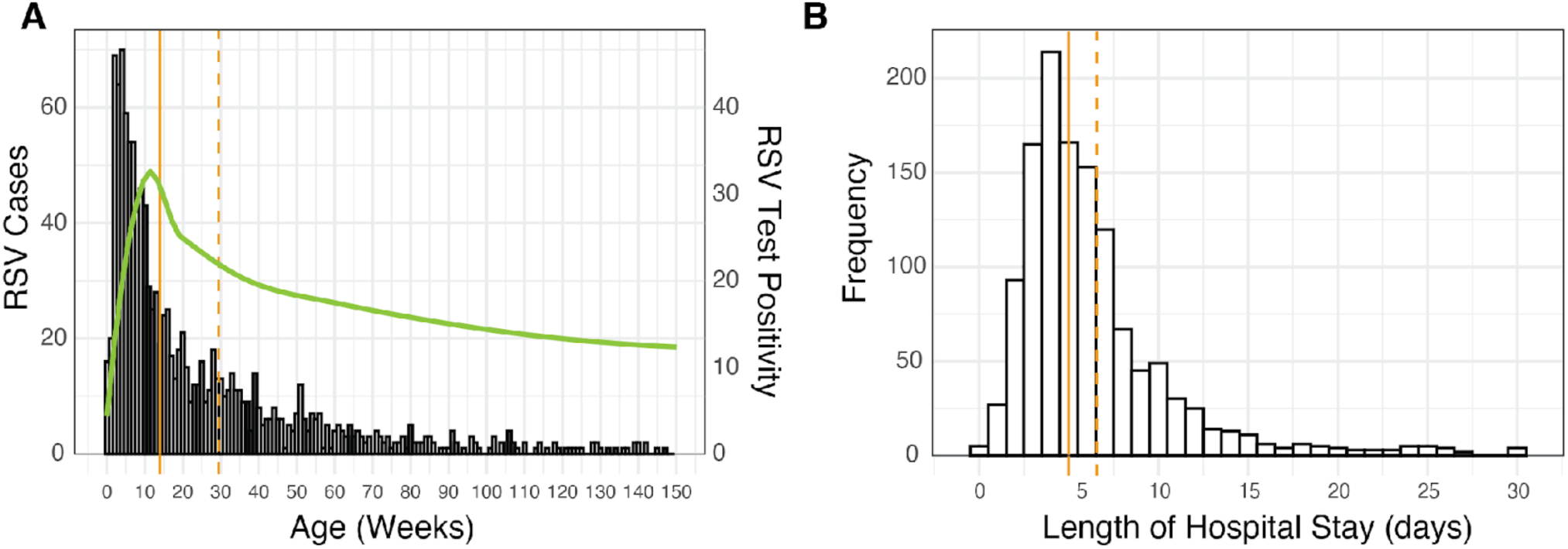
Age distribution of RSV-positive cases and length of hospital stay. A. Age distribution of RSV cases (n = 1,261) in weeks, where the green line represents RSV test positivity in different age groups, the solid orange line indicates the median age, and the dotted orange line shows the mean age. B. Length of hospital stay in days of RSV cases (n = 1,261), with the solid orange line representing the median length of stay and the dotted orange line representing the mean.

### Impact of Bed Shortages on Patients Denied Admission

During the study period, of the total 49,833 children requiring hospitalization, the hospital was unable to admit 9,169 (18.4%) patients due to bed shortages (Figure 1). Of them, we approached and collected contact information of 5,969 families for follow-up; 3,200 patients were not approached because they either were >5 years of age (n=876) or came to the hospital at night when the study research assistants were unavailable (n=2,324) (Figure 1). Of the families who provided contact information for telephone follow-up, a random selection of 3,928 (43%) patients were contacted for follow-up. Out of these, 3,435 (87%) could be successfully contacted, provided verbal consent over telephone to participate and were followed up at 2 weeks (Figure 1). During the first follow-up, 504 (14.7%) were reported to have died. Of the 2,931 patients alive, 2,631 (89.8%) could be subsequently contacted during the 3-month follow-up, and an additional 106 (4%) had died. The survival probability at the end of follow-up was 0.805 (95%CI:0.791-0.819) for denied patients (Figure 4A).

**Figure 4.**
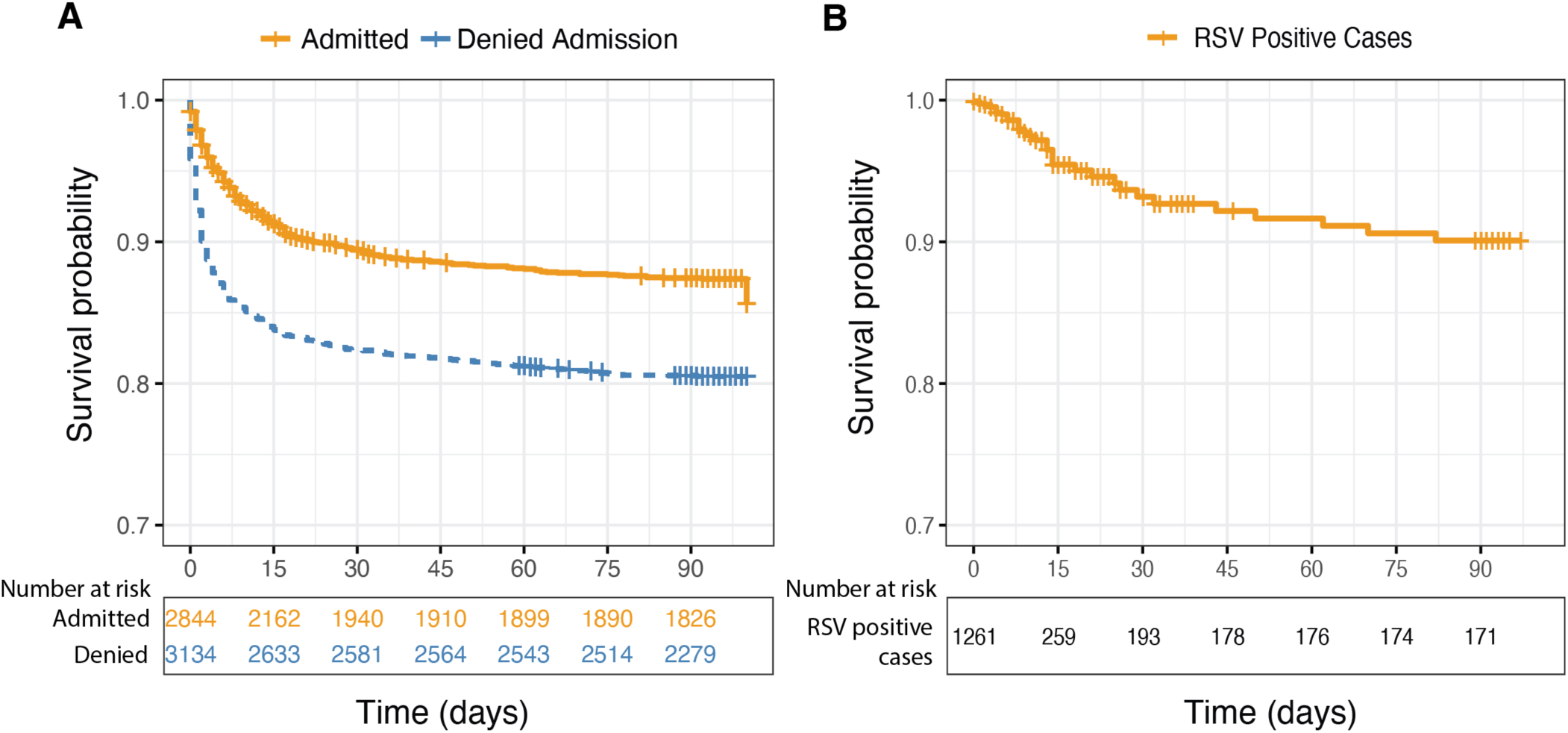
Survival Analysis of Patients followed-up. A. Survival probabilities of patients denied admission and those admitted. Cases were censored at discharge if there was no follow-up or at the last successful follow-up. B. Survival curve for only RSV-positive cases: the 24 cases that resulted in in-hospital deaths, alongside the additional 205 cases that were followed up, of whom 18 subsequently died. Numbers at risk at different time points are provided below the graphs.

2,845 admitted under-five children were randomly selected for follow-up during the same time. Of them, 2,527 (89%) were successfully followed up at 2 weeks over telephone with consent. During the first follow-up, 225 (8.9%) were reported to have died (Table 2). Of the 2,302 patients alive, 1,965 (85.3%) could be subsequently contacted during the 3-month follow-up, among whom an additional 82 (4.2%) had died (Figure 4). The survival probability at the end of follow-up was 0.874 (95%CI:0.861-0.887) among those admitted (Figure 4A).

**Table 2:** Clinical attributes and outcome of participants admitted or denied admissions who were followed-up. The table represents data of participants for whom data could be collected. p-values were computed with Chi-squared tests for categorical variables (sex, diagnosis, mortality), and Kruskal-Wallis test for age. Diagnoses could not be collected for 18% of participants, 14-day mortality was missing for 12% (due to loss to follow u) and 90-day mortality was missing for 21% (due to loss to follow up or mortality at 14 days). *A list of diagnoses that may be included in “other” is listed in the Appendix, Table S1. 90-Day Mortality is among those alive at 14-days, and successfully followed up.

|  | Overall | Admitted | Denied | p-value |
| --- | --- | --- | --- | --- |
| **Number of Patients** | **6772** | **2845** | **3928** |  |
| **Sex - Female (%)** | 2582 (38.1) | 1071 (37.6) | 1511 (38.5) | 0.5077 |
| **Age in months (median [IQR])** | 3.00 [0.00, 12.00] | 4.00 [0.00, 15.00] | 3.00 [0.00, 11.00] | 0.0069 |
| **< 2 weeks** | 2022 (29.9) | 809 (28.4) | 1213 (30.9) | <0.0001 |
| **2 weeks - 3 months** | 1425 (21.1) | 595 (21.1) | 830 (21.1) |  |
| **4 - 6 months** | 622 ( 9.2) | 226 ( 7.9) | 396 (10.1) |  |
| **7 months - 2 yrs** | 1869 (27.6) | 764 (26.9) | 1105 (28.1) |  |
| **2 - 5yrs** | 835 (12.3) | 451 (15.9) | 384 ( 9.8) |  |
| **ER Diagnosis Cluster (%)** |  |  |  |  |
| **Respiratory Manifestation** | 1664 (26.8) | 552 (22.7) | 1112 (29.4) |  |
| **Gastrointestinal Manifestation** | 543 ( 8.7) | 250 (10.3) | 293 ( 7.8) |  |
| **Perinatal Asphyxia** | 772 (12.4) | 281 (11.6) | 491 (13.0) |  |
| **Febrile Illness** | 267 ( 4.3) | 160 ( 6.6) | 107 ( 2.8) | <0.0001 |
| **Systemic Infections** | 577 ( 9.3) | 226 ( 9.3) | 351 ( 9.3) |  |
| **Preterm low-birth weight** | 418 ( 6.7) | 114 ( 4.7) | 304 ( 8.0) |  |
| **Other\*** | 1968 (31.7) | 846 (34.8) | 1122 (29.7) |  |
| **14-day Mortality (%)** | 729 (12.2) | 225 ( 8.9) | 504 (14.7) | <0.0001 |
| **90-day Mortality (%)\*** | 188 (4.1) | 82 (4.2) | 106 (4.0) | 0.8659 |

Overall comparison between the two groups revealed a crude hazard ratio (HR) of 1.69 (95%CI:1.48-1.94) of death in children who were denied admission compared to children who were admitted (Table 3). The HR was 1.56 (95%CI:1.34-1.81) in models adjusted for age, diagnosis (categorized as presented in Table 2), and sex. Models stratified by age groups showed that HR of death was significantly greater for denied cases among babies in their first month of life (HR 2.27 [95%CI:1.87-2.75]) but not in other age groups. When stratified by diagnosis, HR of death was significantly greater among those with perinatal asphyxia (HR 2.05 [95%CI:1.59-2.65]), preterm low-birth-weight (HR 2.78 [95%CI:1.99-3.87]).

**Table 3:**
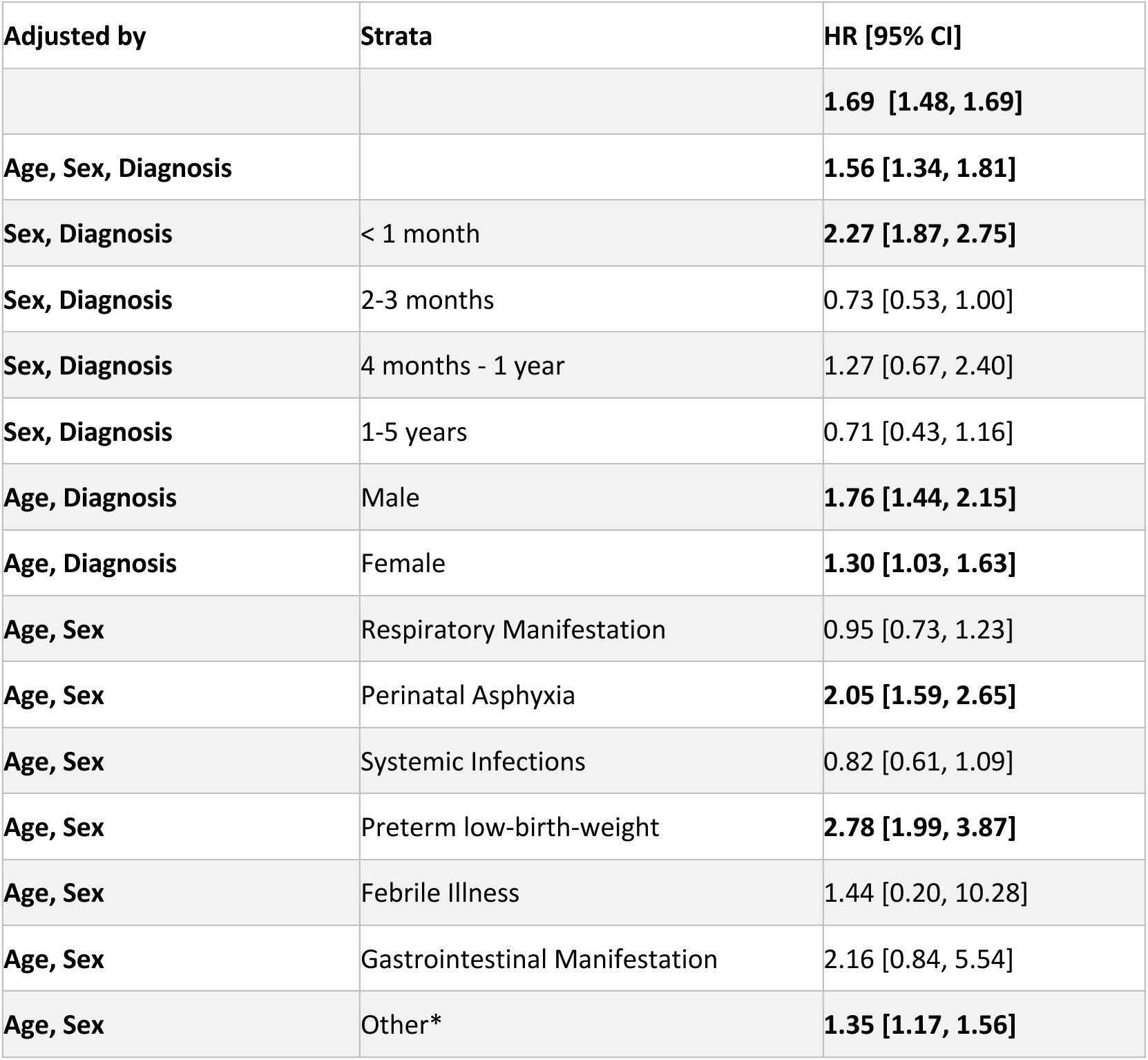
Hazard ratio (HR) of denied vs admitted participants adjusted by age, sex, and diagnosis. *A list of diagnoses that may be included in “other” is listed in the Appendix, Table S1.

| Adjusted by | Strata | HR [95% CI] |
| --- | --- | --- |
|  |  | **1.69 [1.48, 1.69]** |
| **Age, Sex, Diagnosis** |  | **1.56 [1.34, 1.81]** |
| **Sex, Diagnosis** | < 1 month | **2.27 [1.87, 2.75]** |
| **Sex, Diagnosis** | 2-3 months | 0.73 [0.53, 1.00] |
| **Sex, Diagnosis** | 4 months - 1 year | 1.27 [0.67, 2.40] |
| **Sex, Diagnosis** | 1-5 years | 0.71 [0.43, 1.16] |
| **Age, Diagnosis** | Male | **1.76 [1.44, 2.15]** |
| **Age, Diagnosis** | Female | **1.30 [1.03, 1.63]** |
| **Age, Sex** | Respiratory Manifestation | 0.95 [0.73, 1.23] |
| **Age, Sex** | Perinatal Asphyxia | **2.05 [1.59, 2.65]** |
| **Age, Sex** | Systemic Infections | 0.82 [0.61, 1.09] |
| **Age, Sex** | Preterm low-birth-weight | **2.78 [1.99, 3.87]** |
| **Age, Sex** | Febrile Illness | 1.44 [0.20, 10.28] |
| **Age, Sex** | Gastrointestinal Manifestation | 2.16 [0.84, 5.54] |
| **Age, Sex** | Other* | **1.35 [1.17, 1.56]** |

### Mortality among RSV-positive cases

Of the total 1,261 RSV-positive cases recorded in this study, 24 (1.9%) died during their stay at the hospital (Table 1). Median age of these cases was 130 days (IQR:77-175). While the study was not designed to specifically follow up RSV-positive cases, of the 2,527 admitted children randomly selected and successfully followed up, there were 205 RSV-positive cases. Of these, 18 children died within the 90-day follow-up period, six of whom were also recorded as hospital deaths above. This brought the total number of deaths recorded in this study to 36. Details of all cases are provided in the appendix (Table S4). The overall median age was 116.5 days (IQR:69.5-220.3). Considering both in-hospital case fatality of all admissions that tested positive for RSV and mortality during follow-up of a subset of cases, survival decreased substantially over time, with a 90-day survival probability of 0.901 (95%CI:0.865-0.938) at the end of the follow-up period (Figure 4B). The most common final diagnosis was pneumonia/bronchopneumonia (22 cases), frequently associated with additional conditions like sepsis (9 cases), and congenital heart disease (12 cases). RSV cannot be attributed as the underlying cause of death for all these cases, as this study was not designed to establish a causal link between RSV and mortality.

### Potential Impact of RSV preventive interventions

In Monte Carlo simulations of hospital admissions and denials for the baseline scenario, based on the empirical data and a 650-bed capacity, a median of 49,726 children required admissions (95% prediction intervals (PI): 49,355-50,082), and 9,283 (95%PI:8,861-9,714) cases were denied admission over the year. This was in concordance with the true burden of total cases requiring admission and denials – 49,833 and 9,169, respectively. In the scenario where RSVpreF maternal vaccine is introduced with an estimated 38.2% reduction in RSV cases requiring hospitalization (Text S4), 48,972 (95%PI:48,632-49,317) patients required admission, and 8,571 patients were denied admission (95%PI:8,173-8980) – a 1.19 percentage point reduction in median proportion of cases that were denied admissions across simulations (Figure 5A). In the Nirsevimab scenario, with an estimated 69% reduction in RSV cases requiring hospitalization (Text S4), 48,367 (95%PI:48,006 - 48,731) patients required admission, and 7,991 patients were denied admission (95%PI:7,574-8,416) – a 2.2 percentage point reduction (Figure 5A). Figure 5B demonstrates the estimated effects of these interventions on overall mortality. Figures 5C and D compare these interventions with different bed capacities, demonstrating how reducing RSV cases and increasing capacity produce similar effects. Overall, there were 6,970 deaths (95%PI:6832-7117) in the baseline scenario, 6,832 deaths (95%PI:6693-6973) in the RSVpreF vaccination scenario, and 6,727 deaths (95%PI:6576-6862) in the Nirsevimab scenario. This corresponded to an estimated median difference in deaths of 130 (95%PI:-60-322) and 258 (95%PI:32-469), respectively. Supplementary analyses suggested a linear relationship between magnitude of RSV burden reduction and denial rates, holding other parameters of the system constant (Figure S3).

**Figure 5.**
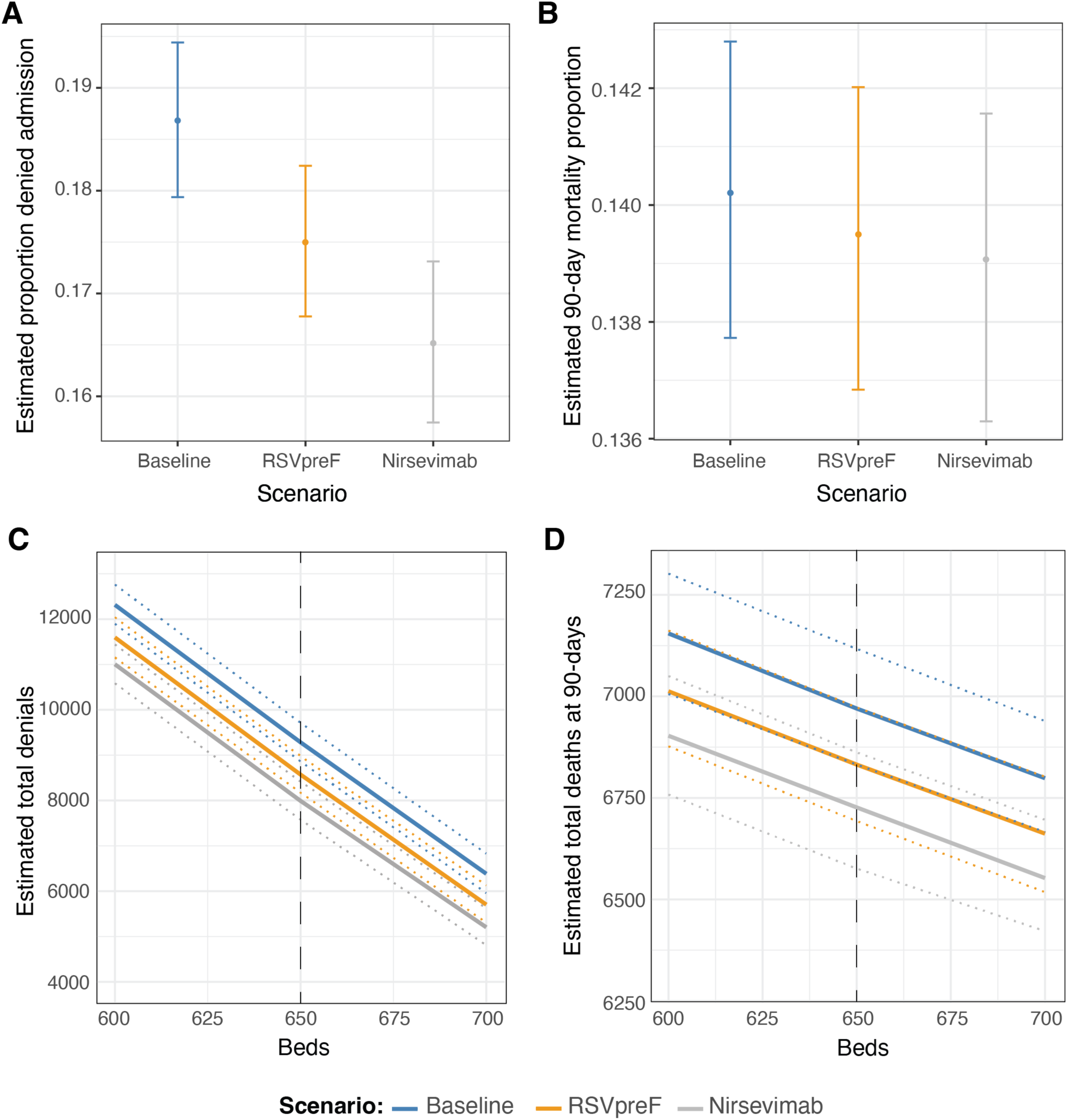
Impact of RSV Preventive Interventions on Hospital Utilization and 90-day Mortality Rates of all children who require admission at BSHI. Effects of a RSVpreF and Nirsevimab on RSV admissions, showing its impact on proportion of hospital denial (A), 90-day mortality proportion (B), total hospital denials (C) and total deaths (D) among children under five. The plots C and D compare mortality against the effective increase in available beds, demonstrating how reducing RSV cases is akin to increasing bed capacity.

## Discussion

Informed decision-making on interventions to prevent RSV infections in young infants requires contemporary, locally relevant epidemiological data. Our study demonstrates RSV’s significant impact on healthcare resources in Bangladesh, a country with a high infectious disease burden and demand for pediatric care amid resource limitations.^12^

Our study recorded a 21% RSV positivity rate among 6,149 patients admitted with respiratory illnesses to Bangladesh’s largest pediatric hospital, corresponding to 465 RSV cases per 10,000 admissions of under-five children. The median age of RSV-confirmed cases was 3 months. These findings align with the PERCH study, which reported 31.2% of children hospitalized with respiratory infections in Bangladesh tested positive for RSV.^14,17^

The in-hospital case fatality for RSV-positive cases was 1.9%. Among 205 randomly selected cases followed for up to 90 days post-hospitalization, 18 deaths were recorded. However, these cases had notable comorbidities, and this study was not designed to attribute these deaths solely to RSV infection. The median age of all recorded RSV-associated deaths was 116 days or 3.8 months. In the Child Health and Mortality Prevention Surveillance (CHAMPS) study, a community-based study not limited to in-hospital deaths, RSV was found in 5.5% of all deaths, with variability across countries: 9.7% in Mali, 10.7% in Ethiopia, and 2% in Bangladesh.^18^ Half of the RSV-associated deaths in the CHAMPS study occurred in infants younger than six months, highlighting the vulnerability of young infants to severe RSV outcomes.

Beyond the direct impact of RSV infections, our findings also demonstrate the considerable burden of infections on the healthcare system. For at least three months during the study year, RSV cases occupied more than 20% of the beds under observation. Median hospital stay for RSV-positive cases was 5 days—longer than the overall median length of stay of 4 days for all admitted patients. In 2019, RSV cases accounted for 8,274 of the total 151,110 bed days observed. Despite being the largest pediatric tertiary hospital with 653 beds, BSHI faces bed shortages often forcing clinicians to deny admission to sick children.^12^ We followed over 5,000 admitted and denied children for up to 90 days to determine the difference in health outcomes and the wider effects of high-burden infections like RSV on the healthcare system. The cumulative mortality proportion for children denied admission was 19.5%, significantly higher than the 12.6% among those admitted. The survival probability at the end of follow-up was 0.874 among those admitted, and 0.805 among those denied admission.

The HR for death was 1.56 among children denied admission compared to those admitted, adjusted for age, diagnosis, and sex. The highest mortality risk was observed in neonates requiring hospitalization within their first month of life, with an HR of 2.27. This disparity highlights the essential role of timely healthcare access in determining child survival outcomes. Monte Carlo simulations of a queueing model helped us characterize hypothetical scenarios with different bed capacities and numbers of patients requiring admission. Were a RSVpreF maternal vaccine or Nirsevimab monoclonal antibody in use, it is estimated to have reduced deaths by 130 and 258, respectively, during the study year among patients requiring admission to the hospital. While the variability of these estimates was large and overlapping, the differences in admission denial rates between scenarios were significant.

Bed shortages are a pressing issue across many LMICs. In Bangladesh, the bed rate of 0.8 per 1,000 population is notably lower than in developed countries, such as 2.9 in the USA and 2.5 in the UK.^19^ With limited infrastructure, preventive strategies are essential. While vaccine introductions and health system improvements over recent decades have reduced under-five mortality in Bangladesh, the rate remains high at 33 deaths per 1,000 live births, highlighting the need for more robust disease prevention.^20^ Given the substantial proportion of beds occupied by RSV cases during peak periods, an RSV maternal vaccine could significantly improve bed availability for critically ill infants. Additionally, administration of monoclonal antibodies, such as Nirsevimab,^3^ in neonates and infants can also be effective, particularly given high preterm birth rates in Bangladesh.^21^ However, cost and logistical challenges complicate the adoption of monoclonal antibody treatments in LMICs like Bangladesh.^22,23^

Global data from various countries have shown shifts in RSV outbreak patterns during and after the COVID-19 pandemic.^24^ While this study was conducted in 2019, before the COVID-19 pandemic, recent reports from Bangladesh suggest that RSV cases have returned to pre-pandemic levels. A report from the Government of Bangladesh highlights a peak in RSV cases among children under five from October 2022 to March 2023, with an average positivity rate of 49% in severe acute respiratory infection cases during that period.^25^ Another study examining RSV-associated deaths from August 2009 to March 2022, found no significant difference in RSV detection rates among deaths before and during the COVID-19 pandemic.^26^

This study has certain limitations to consider. It was conducted in a single hospital, potentially limiting generalizability. Multi-site research could offer a more comprehensive view of the RSV burden across Bangladesh and South Asia. However, as the largest provider of tertiary pediatric care in the country, this hospital’s data is still highly informative. In addition, data collection was limited to one year, meaning that the simulation could only account for RSV’s 2019 seasonal outbreak pattern. Unlike countries in temperate regions, with well-defined RSV seasons, equatorial countries like Bangladesh experience varied RSV peaks—sometimes in winter, summer, or even twice a year—making multi-year data crucial for modeling seasonal variability.^27,28^ Additionally, RSV peaks can overlap with other endemic diseases, such as dengue^29^ and typhoid^30^, which have unpredictable seasonal patterns, complicating generalizable pattern identification even with extended data. The study also did not assess concurrent infections with RSV or confirm deaths directly attributed to RSV. Furthermore, sample attrition and assumptions in the simulation models could introduce bias. Although we achieved a ∼67% follow-up rate for both admitted and denied cases, those lost to follow-up may differ in important ways. During sampling, nasopharyngeal swabs were not collected from children on extensive support, such as high-flow oxygen, possibly underestimating most severe RSV cases with adverse outcomes. In our simulations, we assumed non-study wards had similar seasonal admissions patterns and that overnight denials, not included, were similar to daytime ones. However, children presenting at night, when research assistants were unavailable, may represent a sicker cohort, as emergencies often occur during these hours. This could result in an underestimation of mortality risks associated with hospital denials. As we did not assess care-seeking after denial from BSHI, we cannot disentangle the impact of treatment delay and/or inability to access hospital care. Finally, human factors could have influenced clinicians’ admission decisions during times of bed scarcity even though such decisions should theoretically be unaffected by such constraints.

Despite the limitations, the global health implications of this study are significant. This study underscores the critical role of RSV preventive interventions, such as vaccines and monoclonal antibodies, in reducing the disease burden in resource-limited settings. By preventing RSV-related hospitalizations, vaccination programs could alleviate seasonal surges in bed occupancy, providing much-needed relief to overstretched healthcare systems. Additionally, our findings highlight the broader issue of hospital bed shortages, which remain a significant barrier to effective care in many LMICs. Results support the prioritization of RSV prevention programs and expansion of hospital infrastructure to address persistent capacity challenges, improve healthcare access, and enhance child health outcomes in low-resource settings.

## Author contributions

S.S. contributed to conceptualization, funding acquisition, study design, implementation, monitoring, data analysis, drafting the initial manuscript, and preparation of figures. Su.S. was involved in data analysis, preparation of figures, and manuscript editing. N.K. contributed to data analysis and editing. Y.H. participated in data analysis, preparation of figures, and manuscript editing. M.S.I. was responsible for analysis and coordination of the study, and also contributed to editing. S.I. carried out specimen extraction and PCR testing. Z.B.A. and S.W.R. coordinated and trained doctors and nurses, assisted in the implementation of the study, in addition to contributing to manuscript editing. M.J.A. was involved in monitoring and management activities, as well as editing. A.M.A. performed data cleaning. P.K.S. contributed to management and patient data analysis. M.R.A. was involved in implementation and management. M.R.A. contributed to editing. S.K.S. contributed to conceptualization, funding acquisition, study design, implementation, monitoring, and manuscript editing. All authors reviewed the manuscript and agreed on the contents.

## Data Availability

All data produced in the present work are contained in the manuscript.

## Acknowledgments

We are deeply grateful for the contributions and support of Anannya Barman Jui, Sira Jam Munira, Md. Hafizur Rahman, Shumaiya Ferdaus, Prama Dewan, Ayesha Rahman, Kazi Shammin Azmery Shalda, Shamsun Nahar, Rumana Khan Nitu, Anamika Mazumder, Syeda Mahfuza Zinat, Liza Nath, Tasmim Sultana, Shamima Tarafder, Asyia Akter, Faujiya Khanam Luna, Fatema Akter, Shali Akter, Israt Jahan, Ms. Razia Sultana , Sharmin Jahan, Asheka Shah Munmun, Kaniz Fatema Prity, Md. Shafiqul Islam, Roly Malakar, Shakila Yeasmin and Mahmudul Hasan Sohel. Their collaboration at various stages of the study ensured its successful completion.

## Declaration of Generative AI and AI-assisted technologies in the writing process

During the preparation of this work the authors used ChatGPT to check grammar, spellings, and improve readability of the manuscript. After using this tool/service, the authors reviewed and edited the content as needed and take full responsibility for the content of the publication.

