## appendix for "RSV-Related Healthcare Burden: A Prospective Observational Study in a Resource-Constrained Setting"

*Saha et al*

**Table of Contents**

Supplementary text descriptions of methods 3

Text S1. Enrollment of Cases and Follow-up 3

Text S2. Nasopharyngeal swab collection and qPCR methods 5

Text S3. Survival curve analysis 6

Text S4. Queuing Theory and Monte Carlo Simulation 7

Supplementary Figures 15

Figure S1. Baseline results of daily number of hospital admissions and denials using Monte Carlo simulations. 15

Figure S2. Daily rates of patients requiring hospital admission 15

Figure S3. Various Hypothetical Scenarios of RSV Case Reduction 16

Supplementary tables 18

Table S1. Assigned clusters of related diagnoses. 18

Table S2. Reasons for sample collection failure 24

Table S3. Time of sample collection. 25

Table S4. Details of all recorded deceased RSV-positive cases. 26

Supplementary References 30

### Supplementary text descriptions of methods

#### Text S1. Enrollment of Cases and Follow-up

Patients admitted to the pediatric wards selected for this study were identified using the ward admission logbook. Study physicians conducted a detailed clinical examination and took a thorough medical history. Children aged 0–59 months admitted to the selected wards were evaluated using the RSV hospital-based surveillance case definition.[1] Children hospitalized with a respiratory infection, defined as having cough or shortness of breath, with an onset within the last 10 days, were eligible to be enrolled in the study. In addition, infants younger than six months were also eligible if they presented with apnoea, which is characterized by a temporary cessation of breathing from any cause and/or sepsis (defined as fever (temperature of 37.5 °C or above) or hypothermia (temperature less than 35.5 °C), with indications of shock (lethargy, fast breathing, cold skin, prolonged capillary refill, or a fast, weak pulse). Once a child met the criteria, caregivers were counseled about the study, including its objectives, procedures, and potential risks and benefits. Written consent was mandatory for participation, covering both data collection and nasopharyngeal swab sampling. Following consent, the patient was enrolled, and data was recorded using a standardized, tablet-based system. Clinical follow-up was conducted daily for each enrolled patient until discharge, referral, death, or the family left against medical advice. Physicians and research assistants documented clinical signs, laboratory findings, treatment details, and final outcomes.

For patients denied admission due to bed shortages, research assistants approached the parents upon denial in the ER, and on permission to call them later to follow up on the health status of the child, collected the child’s age, family contact information. Emergency room physician’s diagnosis was also collected. We were very careful about ensuring that caregivers fully understood that the research team would not offer medical assistance. If verbal permission was not provided, no data were collected. This was done as quickly as possible to ensure no delay is caused in the care-seeking process. These families were informed that their participation was optional and would not affect their access to medical care. Selected cases were called via telephone, and verbal consent was taken during these calls.

14 days after admission or refusal, research assistants contacted the caregivers of randomly selected patients by telephone to document the child's health status. Before collecting any health information, the caregiver was reintroduced to the study, including an explanation of the study procedures. Verbal consent for the child's participation in the follow-up study was obtained before collecting any information and documented. Data were only collected for the study if the caregiver provided verbal consent for their participation in the study. Collected details included child’s health status, including symptoms, recovery progress, complications, or hospital readmissions. For children alive on Day 14, an additional follow-up was conducted on Day 90 to assess longer-term health outcomes.

#### Text S2. Nasopharyngeal swab collection and qPCR methods

Nasopharyngeal swab samples were collected by trained nurses using mini flocked swabs (FLOQSwab, Copan Diagnostics, California, USA) and were immediately stored in 1 ml skim milk tryptone-glucose-glycerol (STGG) media. RNA was extracted from 140 µl of the sample volume using the Qiagen QIAamp® Viral RNA Mini kit on QIAcube Connect system (Qiagen, Hilden, Germany). Subsequently, 7 µl of extracted RNA underwent qPCR with qScript XLT 1-Step RT-qPCR ToughMix (QuantaBio, Beverly, MA, USA) and previously reported RSV-specific primers and probe. Final concentrations of primers and probe were 900 nM and 200 nM, respectively, in a 20 µl qPCR reaction volume. qPCR was conducted on Applied Biosystem qPCR 7500 Fast Dx system (Thermo Fisher Scientific, Waltham, MA, USA), with cycling conditions were 50°C for 10 minutes, 95°C for 1 minute, and 40 cycles of 95°C for 10 seconds and 60°C for 1 minute. RSV-positive cases were identified based on true sigmoidal amplification curves and a cycle threshold (Ct) <35.

Forward primer: - 5’- GGCAAATATGGAAACATACGTGAA-3’
reverse primer: 5’-TCTTTTTCTAGGACATTGTAYTGAACAG-3’
probe: 5’-CTGTGTATGTGGAGCCTTCGTGAAGCT-3’ (labeled at the 5’ end with FAM and quenched at the 3’end with Black Hole Quencher-1).

#### Text S3. Survival curve analysis

Survival curves were computed using the Kaplan-Meier method and crude and adjusted hazard ratios were calculated using Cox Proportional-Hazards models. Date of death was collected during follow-up, or from hospital records for admitted cases. Conservative censoring assumptions were made – children denied admission and not followed-up at 14-days were considered censored at day 0, and those not followed up at 90-days were considered censored at 14-days (last successful contact). Admitted children's censoring time was dealt with similarly, except that hospital discharge (either alive or through mortality) was treated as the last available information unless there was further follow-up data. Multivariable Cox Proportional-Hazards models were used to estimate hazard ratios adjusted for diagnosis cluster (Table S1, Appendix page 11), age in months, and sex. We also obtained stratified estimates of hazard ratios.

#### Text S4. Queuing Theory and Monte Carlo Simulation

We used queueing theory and Monte Carlo simulations to understand how a reduction in RSV cases requiring admission may impact strain on hospital capacity, overall refusals of patients seeking admission, and mortality. Queueing theory is used across a wide range of disciplines to understand the dynamics of systems where there is a demand for services, a distribution of service times, and some limitation to service provision that results in wait times or rejection. Queueing models have been in health systems research to optimize resource allocation and system design to minimize patient wait times.[2–4]

In our simulation, the system is initialized with a fixed capacity (number of beds). The average number of patients requiring admission per day is estimated based on the observed pattern of actual daily admissions and denials. The model algorithm progresses day-by-day such that each day:

1) the number of available beds are updated based on discharges;

2) number of patients requiring admission is drawn from a Poisson distribution parameterized using the observed data;

3) all patients requiring admission are admitted unless their number exceed available beds, in which case a random subset is admitted;

4) admitted patients are assigned length-of-stays (and thus discharge dates) drawn from a negative binomial distribution parameterized using the observed data, and

5) the number of available and occupied beds are updated.

Denied and admitted children are assigned separate probabilities of death at 90 days, estimated using the fully adjusted Cox Proportional-Hazards model described above, and standardized to the distribution of covariates in the population. The number of daily denials and admissions were tracked.

Details of parameterization of the algorithm, and a visual depiction is provided below. Different scenarios with various reductions in the number of RSV cases were simulated, including for recently approved maternal vaccine, RSVpreF, and long-acting monoclonal antibodies,

Nirsevimab.[4,5] For the maternal vaccine, we conservatively assumed that no preterm babies (gestational age <37 weeks) will have maternal antibodies, and used a preterm rate of 16.2%. The efficacy used for the licensed RSV maternal vaccine in our population is calculated to be 38.2% and that for the monoclonal antibody to be 69% (full details in the next section below). The median value across 1000 simulation iterations and the 97.5th and 2.5th percentile values across the simulations are reported as the 95% prediction intervals. For percentage denials and percentage mortality (Fig 5A,B), percentages are calculated in each simulation iteration, and the median, 97.5th, and 2.5th percentile values reported as well. For comparing scenarios, simulation iterations were compared across scenarios, and the median, 97.5th and 2.5th percentiles reported.

Details of parameterization of the algorithm, and a visual depiction is provided below.

**Simulation structure and algorithm:**

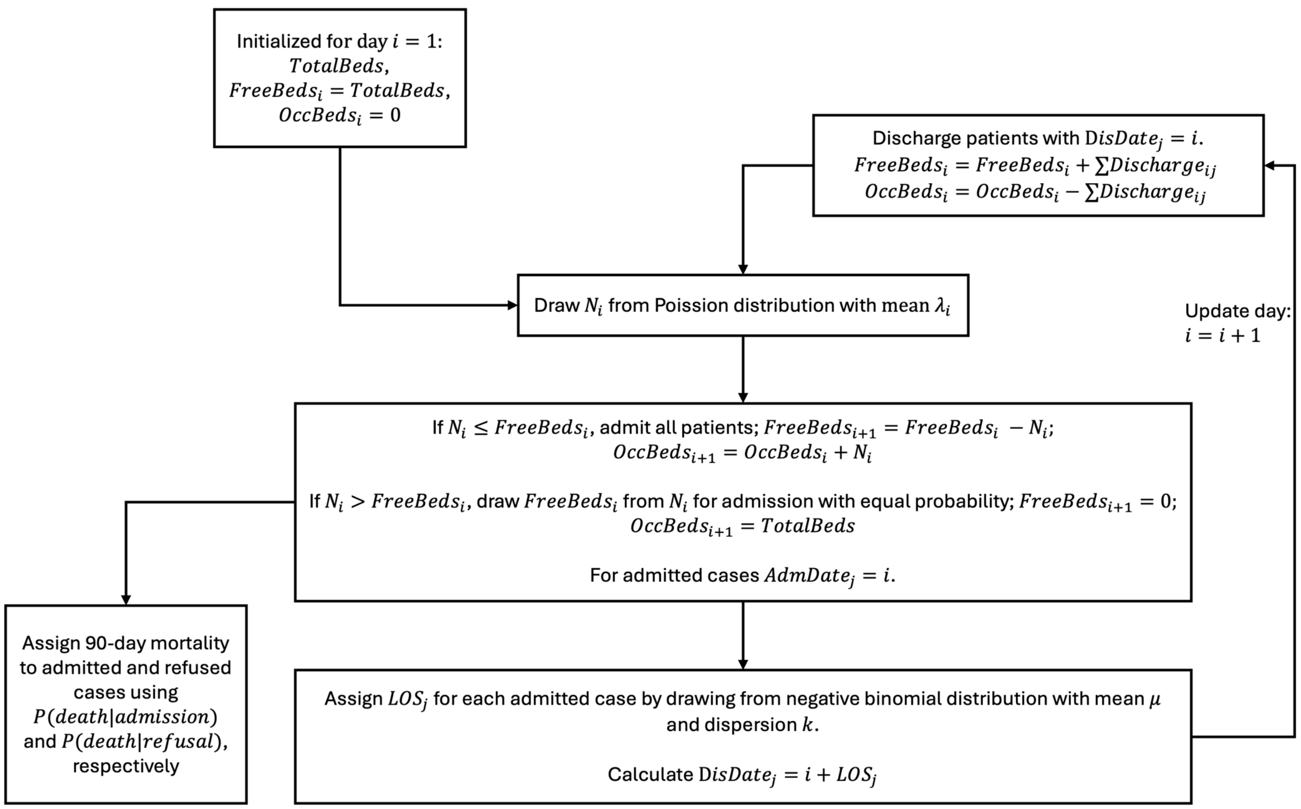

**List of variables and parameters used in the simulation studies:**

| **Variable / Parameter** | **Definition** | **Estimation / Values** |
|---|---|---|
| $TotalBeds$ | The total number of beds available for admissions to the hospital, held fixed throughout the year | Different values set for different scenarios: $TotalBeds = \{450, 500, 550, 600, 650, 700, 800, 850\}$ |
| $FreeBeds_i$ | The total number of unoccupied beds available for patients to be admitted to on a particular day $i$ | Simulated daily based on history of admissions, and discharges based on length-of-stay of patients. |
| $OccBeds_i$ | The total number of occupied beds available on a particular day $i$ | Simulated daily based on history of admissions, and discharges based on length-of-stay of patients. |
| $N_i$ | The total number of patients who arrive at the hospital on a particular day $i$ requiring admission | Drawn from a Poisson distribution where $N_i \sim Poisson(\lambda_i)$ |
| $\lambda_i$ | Rate of patients arriving at hospital requiring admission on a particular day $i$ | Estimated from the daily observed data on admissions and refusals. |

| | | |
|---|---|---|
| | | Total admissions and total refusals for 2019 were known.<br>Daily admissions were known for screening wards. Daily day-time refusals for <5 children were known. To match the known yearly totals and preserve the observed temporal patterns, these daily rates were scaled to match the yearly totals.<br><br>Estimated total daily refusals and admissions were summed and smoothed using a 7-day moving average. These were used as $\lambda_i$ (see figure S2).<br><br>Daily RSV cases were estimated by assuming that:<br>1) the positivity in the eligible and untested children was the same as in the tested children,<br>2) that the non-eligible and non-screened children did not have RSV, and<br>3) that the proportion of all cases that are RSV positive was the same among the admitted and refused cases (since no RSV testing was done among the refused). For different RSV reduction scenarios, $\lambda_i$ was scaled down based on relative reduction of expected RSV cases. |

| | | |
|---|---|---|
| $AdmDate_j$ | Admission date for patient $j$ | Simulated such that $AdmDate_j = i$ if bed is available when a patient requires admission. Further details below |
| $LOS_j$ | Length-of-stay of patient $j$ | Drawn from a negative binomial distribution with mean $\mu$, and dispersion $k$ |
| $\mu, k$ | Mean and dispersion of negative binomial distribution for length-of-stays, respectively | Estimated by fitting a negative binomial distribution to the observed length-of-stays among admitted cases. Estimates were $k = 2.048, \mu = 5.77$ |
| $DisDate_j$ | Discharge date for patient $j$ | Simulated such that $DisDate_j = AdmDate_j + LOS_j$ for admitted patients. Further details below. |
| $Discharge_{ij}$ | An indicator variable for admitted patients: 1 if patient $j$ was discharged on day $i$, 0 if not | Simulated based on Dis$Date_j$. Further details below |
| $Death_j$ | An indicator variable for 90-day mortality: 1 if patient $j$ was alive at 90 days, 0 if not | Bernoulli draw based on admission status and parameters $P(admission), P(death\|refusal)$ |

| | | |
|---|---|---|
| $P(death\|admission)$ $P(death\|refusal)$ | 90-day probability of death for patients who required admission and were admitted or refused, respectively | 90-day survival probability estimated for each group from survival analysis described in Methods. Estimates were: $P(death\|admission) = 0.129$, $P(death\|refusal) = 0.189$ |

**Vaccine Efficacy Calculations**

To estimate the impact of the Pfizer maternal RSV vaccine and monoclonal antibody treatment, we calculated reductions in RSV cases based on age-specific vaccine efficacy data and RSV positivity rates.

1. **RSVpreF vaccine**:

The efficacy of the bivalent vaccine containing pre-fusion F (RSVpreF) varies by age group, according to the findings reported by Kampmann et al.[5] We specifically selected the efficacy data reported for RSV-associated hospitalization.

- <3 months: 67.7%
- 3-6 months: 56.8%
- >6 months: 0%

Age distribution of RSV positive cases in our study:

- <3 months: 55.2%
- 3-6 months: 14.6%
- >6 months: 30.2%

Reduction in RSV cases was calculated using:

$(0.677 \times 0.552) + (0.568 \times 0.146) + (0 \times 0.302) = 0.4566$ or 45.66% reduction.

After accounting for a preterm birth rate of 16.2% in Bangladesh[6] (i.e., mothers unable to receive the vaccine early enough), the overall reduction becomes: $0.4566 \times 0.838 = 0.3826$ or 38.26%.

2. **Nirsevimab**:

The monoclonal antibody Nisevimab has an efficacy of 82.5% for children <1 year of age as reported by Hammitt et al.[7]

Age distribution of RSV positive cases in our study:

- <1 year: 83.7%
- >1 year: 16.3%

Reduction in RSV cases was calculated using:

$0.837 \times 0.825 = 0.6901$ or ~69% reduction.

These reductions were applied to scale down the daily admission rates in the simulation to estimate the impact of RSV case reduction on hospital capacity and patient outcomes.

#### Supplementary Figures

##### Figure S1. Baseline results of daily number of hospital admissions and denials using Monte Carlo simulations.

The circles show actual data from the surveillance; the solid thick lines show the daily median across 100 simulations, and translucent lines show the 100 iterations of the simulation. Note that results in main manuscript are from 1,000 iterations.

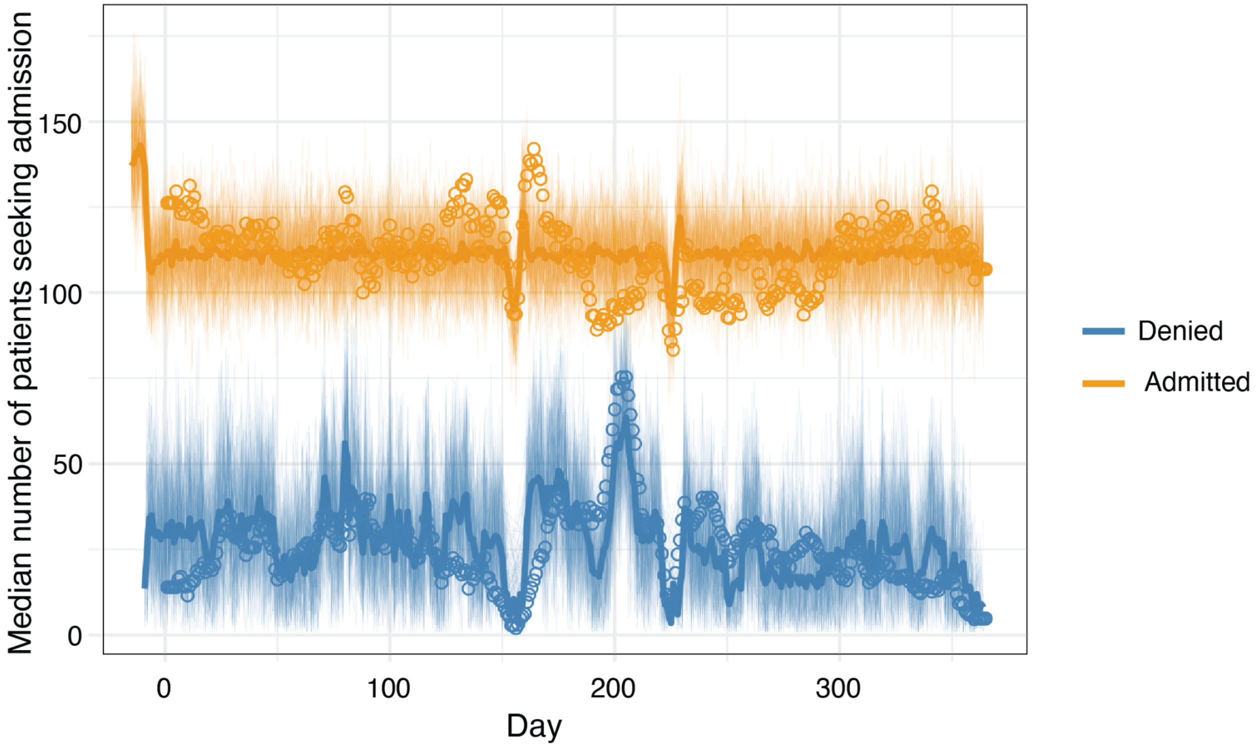

##### Figure S2. Daily rates of patients requiring hospital admission

Seven-day moving averages of the number of patients requiring admission per day in total, for RSV only, and without RSV. The total numbers are the daily means used for Poisson draws in the baseline scenario ($\lambda_i$ ). Hypothetical RSV reduction scenarios use daily means where a proportion of the RSV mean is subtracted from the daily mean.

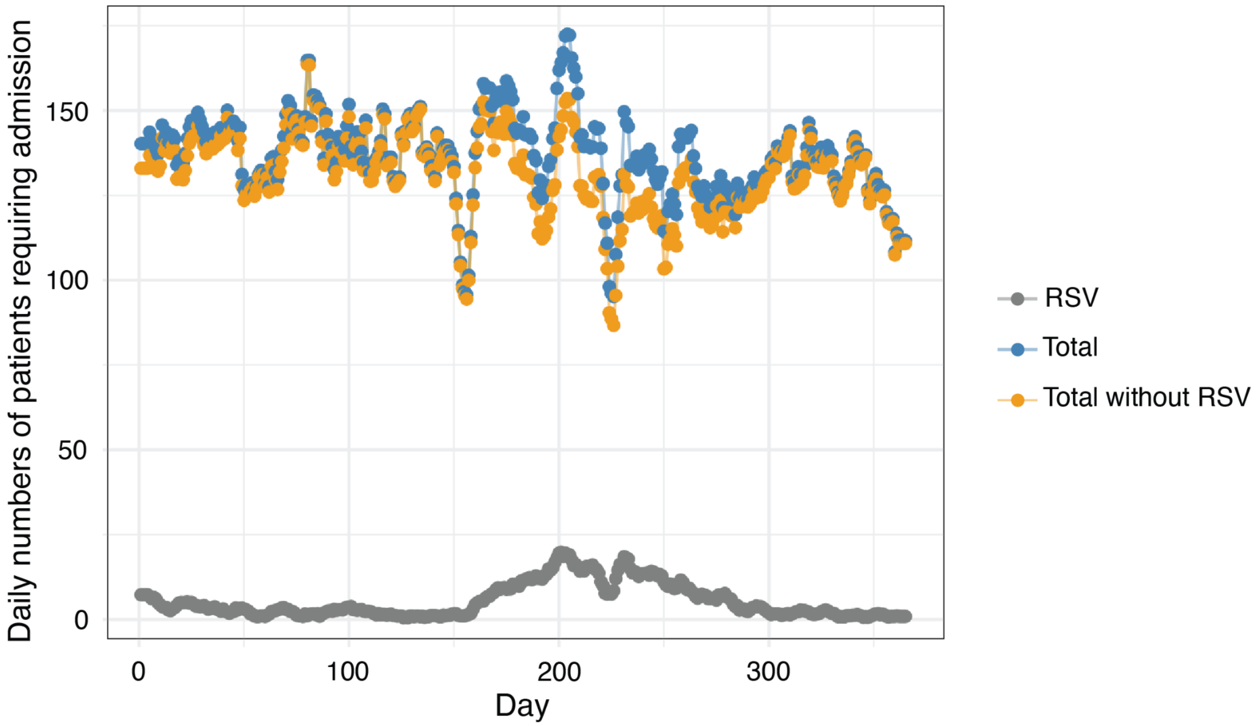

Figure S3. Various Hypothetical Scenarios of RSV Case Reduction

Estimated proportion of admissions that were denied under baseline conditions (0% RSV reduction), and for every 10% additional decrease in RSV burden. Prediction intervals and median for this figure were generated from 100 iterations.

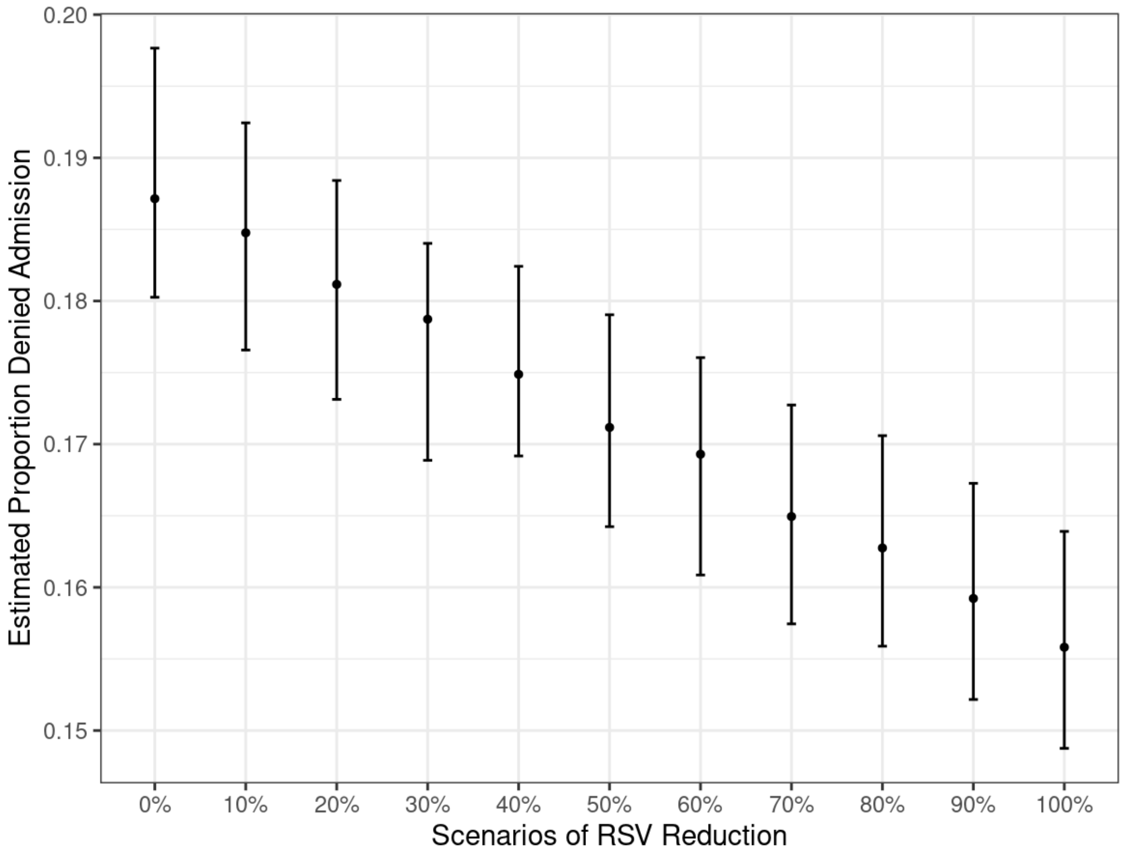

0.20
0.19
0.18
0.17
0.16
0.15
Estimated Proportion Denied Admission
0%
10%
20%
30%
40%
50%
60%
70%
80%
90%
100%
Scenarios of RSV Reduction

#### Supplementary tables

Table S1. Assigned clusters of related diagnoses.

| | |
|---|---|
| **Respiratory manifestation** | Pneumonia/ bronchopneumonia |
| | Severe pneumonia |
| | Bronchiolitis/ acute bronchiolitis |
| | ARI/Acute respiratory infection |
| | RDS/ Respiratory distress Syndrome |
| **Systemic infections** | Septicaemia / sepsis |
| | Neonatal sepsis |
| **Febrile illness** | Enteric fever/ Typhoid fever / Paratyphoid fever |
| | Febrile convulsion / Atypical febrile convulsion |
| | Viral fever/Dengue Fever |
| **Neurological manifestation** | Meningitis |
| | Encephalitis/Meningoencephalitis/Encephalopathy |
| | Seizure disorder/Neonatal seizure |
| | Epilepsy |
| **Gastrointestinal manifestation** | Acute gastroenteritis/ AGE/Acute watery diarrhea |
| | Persistent diarrhoea |
| | Dysentery/ Shigellosis/ Invasive diarrhoea |

| | |
|---|---|
| **Genitourinary/Renal manifestation** | Nephrotic syndrome |
| | Renal failure/ kidney failure |
| | AGN/ Acute Glomerulonephritis/ APSGN |
| **Cardiovascular manifestation** | Ventricular septal defect/ VSD |
| | Tetralogy of Fallot/ TOF |
| | Other congenital heart disease |
| **Perinatal asphyxia** | Perinatal asphyxia |
| **Preterm low-birth weight** | Preterm low-birth weight |
| **Neonatal jaundice** | Neonatal jaundice |
| **Other** | Other* |

***Other may include the following diagnoses:**

| |
|---|
| Neonatal jaundice |
| Neurological manifestation |
| Genitourinary/Renal manifestation |
| Congenital Cardiovascular manifestation |
| Acute abdomen |
| ABO incompatibility/ Rh incompatibility |
| Achondroplasia |
| Acute Appendicitis/ burst appendix |
| Acute Epididymoorchitis/ Orchitis |
| Acute flaccid paralysis/ AFP |
| Acute gastritis |
| Acute laryngotracheobronchitis |

| |
|---|
| Club foot/ TEV |
| CMV infection |
| Collodion baby |
| Congenital anomaly/ Cloacal anomaly |
| Congenital pneumonia |
| Congenital rubella syndrom |
| Congenital Syphillis |
| Constipation |
| Cough & cold/common cold |
| Croup |
| Cystic fibrosis |
| Cystic hygroma |

| |
|---|
| Acute leukemia (ALL/AML) |
| Acute otitis media/Otitis media (ASOM) |
| Acute scrotum |
| Acute Severe Asthma/Acute exacerbation |
| Acute stroke syndrome |
| Adrenal Hyperplasia |
| AEFI/ Adverse effect following immunization |
| Anaemia |
| Anal fissure<br>Anorectal malformation/ ARM |
| Anuria/ oliguria |
| Aplastic anemia |
| Arthritis/ rheumatoid arthritis/ JIA |
| Ascariasis/ helminthiasis |
| Aspiration Pneumonia |
| Aspiration syndrome |
| Ataxia/ cerebeller ataxia |
| Bell's palsy/ facial nerve palsy |
| Biliary atresia |
| Birth injury |
| Bladdar Obstruction/ urinary obstruction |
| Bleeding disorder |
| Breath holding attack |
| bullous disorder/ Epidermolysis Bullosa |
| Burn/ scald/ post burn contracture |
| Cardiomyopathy/ cardiomegaly |
| Cellulitis |
| Cerebral atrophy/ cortical atrophy |

| |
|---|
| Dengue fever |
| Dermatitis |
| Developmental delay |
| Diabetes/ Diabetes mellitus |
| Down's syndrome |
| Drug overdose/drug reaction/poisoning |
| Electrolyte imbalance |
| Empyema thoracis |
| Encephalopathy |
| Eventration of diaphragm |
| Extrapulmonary Tuberculosis |
| Failure to thrive/FTT |
| Feeding problem/feeding mismanagement |
| Fistula |
| G-6-PD Deficiency |
| Gastro esophageal reflux disease/ GERD |
| Growing pain |
| Guillain- Barre syndrome/GBS |
| Gut atresia/ coanal atresia/ intestinal |
| Gut malrotation |
| Haemorregic diseases of new born (HDN) |
| HB-E Diseases |
| Head injury |
| Healthy baby/ Normal baby/ well baby |
| Heart failure |
| Hemangioma |
| Hematemesis |
| Hematoma/ caphal hematoma |

| |
|---|
| Cerebral malaria/malaria/severe malaria |
| Cerebral palsy |
| Chicken pox |
| Chocking/ chocking Attack |
| Cholecystitis/ choledocal cyst |
| Chronic kidney disease/ CKD |
| Chronic suppurative otitis media (CSOM) |
| CLD/chronic liver disease |
| Cleft lip/Cleft palate |
| Hydrocephalus |
| Hydronephrosis |
| Hypoglycemia |
| Hypospadiasis |
| Hypothyroidism |
| ICSOL/ Intracranial space occupying lessions |
| Icthyosis |
| IDM/ infant of diabetic mother |
| Infantile colic/ infantile spasm |
| Infantile hypertrophic pyloric stenosis |
| Infected scabies/Scabies |
| Infective endocarditis |
| Injury/ Cut injury/ lacerated injury |
| Insect bite/ bee bite |
| Intestinal obstruction/ paralytic ileus |
| Intestinal perforation |
| Intestinal Tuberculosis |
| Intrauterine Growth retardation (IUGR) |
| Intussusception |

| |
|---|
| Hemiplegia |
| Hemolytic anemia |
| Hepatitis/neonatal hepatitis Syndrome |
| Hepatoblastoma |
| Hernia/ Inguinal hernia |
| HIE/ Hypoxic-ischaemic encephalopathy |
| Hirschsprung disease/ HD |
| HTN/ Hypertension |
| Hydrocele/ Encysted hydrocele |
| Per rectal bleeding |
| Pericardial effusion |
| Pharyngitis |
| Phymosis/ para phymosis |
| Pierre robin syndrom |
| Pleural effusion |
| Pneumonic consolidation |
| Pneumonitis |
| Pneumothorax/Hydro pneumothorax |
| Polycystic kidney disease/ PKD |
| Portal HTN/ portal hypertension |
| Post circumcission bleeding |
| Post measles pneumonia |
| Post meningitis neurological sequale |
| Postdated baby |
| Pseudotumor cerebri |
| Pulmonary hypertension/ PAH |
| Pure red cell aplasia |
| PUV/ posterior urethral valve |

| |
|---|
| ITP /Idiopathic Thrombocytopanic Purpura |
| Jaundice |
| Jejunal atresia |
| Kala azar |
| Laryngomalacia |
| Lipoma |
| Liver abscess/ Amoebic liver abscess |
| Liver failure/ hepatic failure |
| Low Birth weight (LBW) |
| Lung abscess |
| Lung collapse |
| Lymphadenitis/ lymphadenopathy |
| Lymphoma/ Hodgkin's disease |
| Measles |
| Meconium aspiration syndrome (MAS) |
| Meduloblastoma |
| Melingaring |
| Meningocele/ myelocele/ meningomyocele |
| Meningoencephalitis |
| Microcephaly |
| Motor neuron disease/ UMN |
| Mumps |
| Muscular atrophy/ muscular dystrophy |
| Myopathy/ Myositis/ myalgia |
| Near drowning |
| Neonatal convulsion/neonatal seizure |
| Nephroblastoma |
| Neuroblastoma |

| |
|---|
| Pyrexia of unknown origin (PUO) |
| Rabies |
| Reactive airway disease |
| Rectal polyp |
| Respiratory wheeze/recurrent wheeze |
| Rheumatic fever |
| Rickets |
| Ricketsial fever |
| Scarlet fever |
| Septic arthritis |
| Sequelae |
| Some/severe dehydration |
| Stevens Johnson Syndrome |
| Stomatitis/ Gingivo Stomatitis |
| Storage disease |
| Sub dural effusion |
| Subacute sclerosing panencephalitis |
| Synovitis/ transient synovitis |
| Teratoma |
| TGA/ Transposition of great artery |
| Thalassemia |
| Tonsilitis |
| TORCH infection |
| TTN/ transient tachypnea of newborn |
| Tubercular meningitis |
| Tuberculosis/disseminated tuberculosis |
| Umbilical Hernia |
| Umbilical sepsis |

| |
|---|
| Neurodegenerative disorder |
| Neurometabolic disease |
| Omphalitis |
| Omphalocele |
| Oral Thrush |
| Osteomyelitis/ Acute Osteomyelitis |
| Pancreatitis/ Acute pancreatitis/ Chron |
| PEM/2° PEM |
| Peptic ulcer disease/ PUD |

| |
|---|
| Undescendent testis/ UDT |
| Upper respiratory tract infection (URTI) |
| Urinary tract infection (UTI) |
| Urine suppression |
| Urticaria/ allergic reaction |
| Valvular heart disease |
| Whopping cough |
| Wilms tumor |

Table S2. Reasons for sample collection failure

| Reasons | Number of Cases | Percentage |
|---|---|---|
| Did not consent | 152 | 14.6 |
| Use of high flow oxygen canula, Cpap/incubators/headbox/intubation | 133 | 12.8 |
| Moved to a special ward | 64 | 6.1 |
| Discharged before specimen collection | 669 | 60.1 |
| Other reasons | 24 | 6.5 |

Table S3. Time of sample collection.

Time of sample collection was available for 6119 of 6149 samples.

| **The interval between time of admission and sample collection** | **Number of samples collected (%)** | **Number of RSV-positive samples collected (%)** |
|---|---:|---:|
| 0 - 23 hours | 2,732 (44.6%) | 537 (42.7%) |
| 24 - 47 hours | 2,161 (35.3%) | 467 (37.1%) |
| 48 - 71 hours | 717 (11.7%) | 155 (12.3%) |
| >72 hours | 509 (8.3%) | 99 (7.9%) |

Table S4. Details of all recorded deceased RSV-positive cases.

| ID | Sex | Age (range in days) | Hospital duration (days) | Hospital Outcome | Selected for follow up | Days between death and admission | Place of death | Final Diagnoses at study hospital |
|---|---|---|---|---|---|---|---|---|
| 1 | F | 91-180 | 0 | Died | Yes | 0 | Study hospital | Acute respiratory infection/Respiratory tract infection, Heart failure, PEM/Kwashiorkor/Marasmus/Severe malnutrition/Oedematous malnutrition |
| 2 | M | 91-180 | 1 | Died | No | 1 | Study hospital | Cholecystitis/ choledocal cyst/ cholelithiasis, Liver failure/ hepatic failure |
| 3 | F | 181-365 | 2 | Died | No | 2 | Study hospital | Pneumonia/bronchopneumonia, HTN/Hypertension/essential hypertension |
| 4 | M | 91-180 | 3 | Died | Yes | 3 | Study hospital | Pneumonia/bronchopneumonia |
| 5 | M | 91-180 | 3 | Died | No | 3 | Study hospital | Septicaemia/sepsis, Pneumonia/bronchopneumonia, Meningitis |
| 6 | F | 31-90 | 4 | Died | Yes | 4 | Study hospital | Pneumonia/bronchopneumonia, Septicaemia/sepsis, Failure to thrive/FTT |

| | | | | | | | | |
|---|---|---|---|---|---|---|---|---|
| 7 | F | 91-180 | 4 | Died | No | 4 | Study hospital | Septicaemia/sepsis, Other |
| 8 | M | 31-90 | 4 | Died | No | 4 | Study hospital | Severe pneumonia, Heart failure |
| 9 | F | 181-365 | 4 | Died | No | 4 | Study hospital | Pneumonia/bronchopneumonia, Septicaemia/sepsis |
| 10 | F | 31-90 | 5 | Died | No | 5 | Study hospital | Pneumonia/bronchopneumonia, Other congenital heart disease (PDA/ASD), Heart failure |
| 11 | M | 0-30 | 6 | Died | No | 6 | Study hospital | Other congenital heart disease |
| 12 | F | 91-180 | 6 | Died | No | 6 | Study hospital | Hernia/ Inguinal hernia/ostructed hernia/ incisional hernia/strangulated hernia |
| 13 | F | 91-180 | 8 | Died | No | 8 | Study hospital | Pneumonia/bronchopneumonia, Ventricular septal defect, Other congenital heart disease |
| 14 | F | 91-180 | 8 | LAMA | Yes | 8 | Road | Pneumonia/bronchopneumonia |
| 15 | F | 0-30 | 8 | Died | Yes | 8 | Study hospital | Severe perinatal asphyxia/perinatal asphyxia, HIE, Neonatal jaundice, Neonatal sepsis |
| 16 | M | 91-180 | 9 | Died | Yes | 9 | Study hospital | Pneumonia/bronchopneumonia, Septicaemia/sepsis, Failure to thrive |

| | | | | | | | | |
|---|---|---|---|---|---|---|---|---|
| 17 | M | 31-90 | 10 | Died | Yes | 10 | Study hospital | Pneumonia/bronchopneumonia |
| 18 | M | 0-30 | 11 | Died | No | 11 | Study hospital | Ventricular septal defect, congenital heart disease, Cardiomyopathy/cardiomegaly |
| 19 | M | 181-365 | 13 | Died | No | 13 | Study hospital | Seizure disorder |
| 20 | F | 366+ | 13 | Died | No | 13 | Study hospital | Pneumonia/bronchopneumonia, Developmental delay |
| 21 | M | 91-180 | 14 | Died | No | 14 | Study hospital | Pneumonia/bronchopneumonia |
| 22 | F | 91-180 | 14 | Died | No | 14 | Study hospital | Pneumonia/bronchopneumonia, Septicaemia/sepsis, Intestinal obstruction/ paralytic ileus/ Gastric Outlet obstruction |
| 23 | F | 181-365 | 14 | Died | No | 14 | Study hospital | Pneumonia/bronchopneumonia, Ventricular septal defect, Down's syndrome |
| 24 | F | 0-30 | 25 | Died | No | 25 | Study hospital | Severe perinatal asphyxia/perinatal asphyxia, Neonatal sepsis, Neonatal jaundice, Other congenital heart disease |
| 25 | M | 366+ | 26 | Died | No | 26 | Study hospital | Pneumonia/bronchopneumonia, Septicaemia/sepsis, Developmental delay, Pleural effusion |

| | | | | | | | | |
|---|---|---|---|---|---|---|---|---|
| 26 | M | 181-365 | 4 | LAMA | Yes | 6 | Home | Pneumonia/bronchopneumonia |
| 27 | M | 91-180 | 8 | LAMA | Yes | 18 | Home | Pneumonia/bronchopneumonia |
| 28 | M | 91-180 | 11 | Discharge | Yes | 21 | Home | Pneumonia/bronchopneumonia, Laryngomalacia |
| 29 | M | 0-30 | 7 | Discharge | Yes | 29 | Home | HTN/ Hypertension/ essential hypertension, Neonatal sepsis |
| 30 | F | 181-365 | 7 | Discharge | Yes | 32 | Hospital | Ventricular septal defect (VSD) |
| 31 | M | 31-90 | 6 | LAMA | Yes | 32 | Home | Pneumonia/bronchopneumonia, Other congenital heart disease |
| 32 | M | 0-30 | 4 | Discharge | Yes | 43 | Hospital | Pneumonia/bronchopneumonia, Ventricular septal defect, Other congenital heart disease, Transposition of great artery/dextro-Transposition |
| 33 | M | 91-180 | 5 | LAMA | Yes | 50 | Home | Pneumonia/bronchopneumonia, Other congenital heart disease |
| 34 | F | 0-30 | 7 | Discharge | Yes | 62 | Hospital | Pneumonia/bronchopneumonia, Other congenital heart disease |

| | | | | | | | | |
|---|---|---|---|---|---|---|---|---|
| 35 | M | 181-365 | 6 | Discharge | Yes | 70 | Home | Pneumonia/bronchopneumonia, Other congenital heart disease, Ventricular septal defect, Down's syndrome |
| 36 | F | 366+ | 10 | Discharge | Yes | 82 | Home | Pneumonic consolidation, Pleural effusion |
